# The safety and relative effectiveness of non-psychoactive cannabinoid formulations for the improvement of sleep: a double-blinded, randomized controlled trial

**DOI:** 10.1101/2023.01.20.23284842

**Authors:** Jessica Londeree Saleska, Corey Bryant, Antonija Kolobaric, Christopher S. Colwell, Derek Loewy, Jeff Chen, Emily K. Pauli

## Abstract

The objective of this randomized, double-blinded controlled trial was to evaluate the safety and relative effects of different formulations containing Cannabidiol (CBD) and melatonin, with and without the addition of minor cannabinoids, on sleep. Participants (N=1,793 adults experiencing symptoms of sleep disturbance) were assigned to receive a 4-week supply of 1 of 6 products (all capsules) containing either 15mg CBD or 5mg melatonin, alone or in combination with minor cannabinoids. Sleep disturbance was assessed using Patient-Reported Outcomes Measurement Information System (PROMIS™) Sleep Disturbance SF 8A, administered via weekly online surveys. All formulations exhibited a favorable safety profile (12% of participants reported a side effect and none were severe) and led to significant improvements in sleep disturbance (p<0.001 in within-group comparisons). Most participants (56% to 75%) across all formulations experienced a clinically important improvement in their sleep quality. There were no significant differences in effect, however, between 15mg CBD isolate and formulations containing 15mg CBD and 15mg Cannabinol (CBN), alone or in combination with 5 mg Cannabichromene (CBC). There were also no significant differences in effect between 15mg CBD isolate and formulations containing 5 mg melatonin, alone or in combination with 15mg CBD and 15mg CBN. Our findings suggest that chronic use of a low dose of CBD is safe and could improve sleep quality, though these effects do not exceed that of 5 mg melatonin. Moreover, the addition of low doses of CBN and CBC may not improve the effect of formulations containing CBD or melatonin isolate.

## INTRODUCTION

Approximately one third of American adults do not get enough sleep each night.^1^ Poor sleep can have a profound impact on a person’s quality of life; it can hinder cognitive functioning^2^ and lead to depression,^3^ reduced productivity,^4^ cardiovascular disease,^5^ and increased healthcare utilization.^4^ There is strong clinical evidence in support of pharmacologic interventions for the treatment of insomnia (difficulty getting to sleep or staying asleep^6^) and other sleep disorders, in particular for GABA_A_ receptor agonists, such as benzodiazepines.^7^ Concerns remain, however, over their many side effects, including ‘hangover effects’, cognitive impairment, abuse, and the considerable risk of dependance.^8^ Consequently, there is a prevailing need to evaluate safer forms of therapeutic treatment for the improvement of sleep.

Many patients turn to complementary and alternative medicines (CAM) for the treatment of insomnia and other sleep disorders.^9^ Melatonin is among the most commonly used and well-studied CAM treatments for sleep,^10^ and clinical evidence supports its efficacy for the improvement of sleep quality, particularly for those experiencing jet lag and delayed sleep-wake phase disorder.^11–14^ Moreover, melatonin exhibits a favorable safety profile and does not demonstrate dependence even when administered at high doses.^15^

Cannabis preparations have also begun to gain attention for their potential therapeutic effects for the treatment of insomnia and other sleep disorders.^16^ To date, the preponderance of clinical research on Cannabis and sleep has focused on Δ9-tetrahydrocannabinol (Δ9-THC), the major active constituent of Cannabis sativa.^17^ Yet use of the non-psychoactive cannabinoid Cannabidiol (CBD) has proliferated in the US, with many new users seeking relief for sleep difficulties.^18^ Preclinical research has demonstrated that CBD possesses anxiolytic, anti-inflammatory, and analgesic properties,^19^ which could aid in the improvement of sleep. Evidence from retrospective and prospective observational studies also suggest that the clinical administration of cannabinoids could improve sleep and other related health issues such as pain and anxiety.^20–22^

Clinical research assessing the use of CBD for insomnia and other sleep disorders remains limited, though some small clinical studies have found support for the hypothesis that CBD may improve sleep. In a study of 15 individuals with insomnia, those who received 160 mg CBD reported sleeping longer than those who received placebo.^23^ Another study of 33 individuals with Parkinson’s Disease revealed that 300 mg of CBD per day led to a transient improvement in sleep quality relative to placebo.^24^ In small experimental studies, fixed doses of 300 mg, 400 mg and 600 mg of CBD were also found to induce self-reported sedative effects relative to placebo in healthy adults (11 adults, 300 and 600 mg^25^; 10 males, 400 mg^26^). Importantly, clinical evidence of CBD also indicates that the cannabinoid has a favorable safety profile,^27,28^ even when taken at doses as high as 1200 mg daily for up to 4 weeks,^29^ supporting the exploration of CBD as a potentially safer therapeutic option for the improvement of sleep.

Despite the limited clinical evidence, marketing claims regarding the effectiveness of CBD for sleep abound.^30,31^ Many manufacturers have also touted the superiority of their CBD products relative to melatonin^32,33^ though, to date, no clinical study has directly compared the effects of these compounds on sleep. Manufacturers have also combined CBD with melatonin and other minor cannabinoids, claiming that these additions could enhance the effect of CBD or melatonin alone.^34^ These claims, too, are unfounded. No large scale randomized clinical trial has evaluated whether CBD could impact the effects of melatonin on sleep (or vice versa). No clinical trials have also evaluated whether the addition of minor cannabinoids, such as Cannabichrome (CBC) and Cannabinol (CBN), could contribute to the therapeutic effectiveness of CBD for sleep improvement. CBN, in particular, has gained prominence as a sleep aid additive,^35^ though the literature is almost entirely devoid of clinical research supporting its effect on sleep quality.^36^ Multi-arm studies allowing for direct comparisons across cannabinoid and melatonin formulations could provide critical information on the main and interactive effects of these compounds on sleep.

Notably, few clinical trials of CBD have also tested the effect of daily usage of CBD at the lower doses commonly found within commercially available products. Most clinical trials of CBD evaluate doses ranging from 300 to 1500 mg CBD per day,^37–39^ while commercial products’ dosage generally range from 5 to 100 mg CBD per day. Clinical research on CBD has suggested a bell-shaped dose response curve wherein an intermediate dose of CBD exhibits a greater anxiolytic effect than a very low or high dose,^40,41^ though more clinical studies are needed evaluate the therapeutic benefits of chronic CBD use for sleep at dose ranges reflecting that of commercial products.

This study sought to address these gaps in the literature by investigating the effect of chronic use (daily use over 4 weeks) of low dose CBD and melatonin formulations, alone and in combination with certain minor cannabinoids, on sleep quality. The primary objective of the study was to compare the safety and effects of CBD isolate to CBD combination formulations (i.e., formulations containing CBD and minor cannabinoids, with and without melatonin) to determine if the addition of melatonin and these minor cannabinoids confer any therapeutic benefit to a formulation containing CBD. As a secondary aim, we also sought to determine if the addition of CBD and CBN confer any therapeutic benefit to a formulation containing melatonin.

## MATERIALS AND METHODS

### Study design and participants

This study, referred to as Radicle Discovery™ Sleep, was a randomized, parallel design, double-blinded controlled study. Radicle Discovery™ Sleep was entirely virtual; no in-person visits were required as data were collected via online surveys and communication with participants occurred through email or SMS text message.

Participants were recruited online from across the US through social media, Radicle Science’s electronic mailing list, and a third-party consumer network with nationwide representation. Recruitment emails containing links to the study screener were sent to those within the Radicle Science mailing list and consumer network, while social media advertisements provided direct links to the study screener. Participants were eligible if they were 21 years old or older, resided in the United States and reported that they were experiencing symptoms of sleep disturbance. Individuals were excluded if they were pregnant or breastfeeding or taking medications with which cannabinoids could interfere. Eligible individuals we directed to a secure online portal to provide informed consent. Participants indicating their consent could electronically sign and date the informed consent form and were sent a digital copy of the electronic consent. Eligible individuals were advised to consult with their healthcare provider before participating if they had a diagnosed medical condition, were on any prescription medication or supplements, or had any upcoming medical procedures planned. Immediately following informed consent, participants then completed an intake survey which collected basic demographic information, health behaviors, and sleep quality.

Those who consented to participate and completed intake were then randomized to one of six study arms (see below for details on randomization). There were 6 arms total within the study; 2 isolate formulations (15 mg CBD; 5 mg melatonin), and 4 combination formulations (all of which contained non-psychoactive cannabinoids; see **Table 1**). All were capsules. Participants were sent a 4-week supply of their study product in the mail along with instructions for study participation within the product insert. All products were provided by the partnering manufacturer, Open Book Extracts, and analyzed at an independent laboratory to ensure active ingredient identification, safety and potency. Participants were informed that they could escalate their dosage from 1 capsule per day to a maximum of 2 capsules per day as needed throughout the study. See **Table 1** for more detail on the formulation and usage of each study product arm. The study was double-blind; neither the participants nor those who collected the data were aware of the product participants received until the conclusion of the study.

**TABLE 1:**
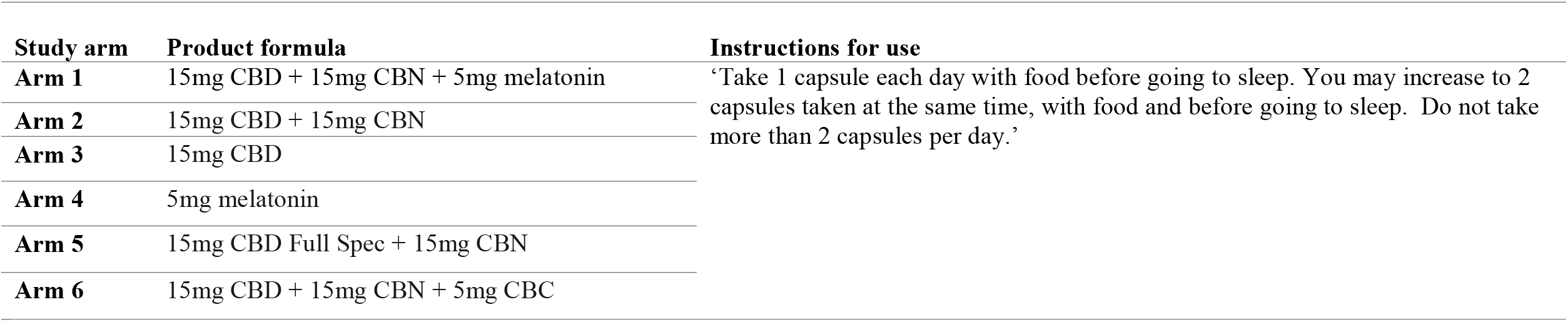
Study arms and instructions for use in the Radicle Discovery™ Sleep study.

| Study arm | Product formula | Instructions for use |
| --- | --- | --- |
| Arm 1 | 15mg CBD + 15mg CBN + 5mg melatonin | ‘Take 1 capsule each day with food before going to sleep. You may increase to 2 capsules taken at the same time, with food and before going to sleep. Do not take more than 2 capsules per day.’ |
| Arm 2 | 15mg CBD + 15mg CBN |  |
| Arm 3 | 15mg CBD |  |
| Arm 4 | 5mg melatonin |  |
| Arm 5 | 15mg CBD Full Spec + 15mg CBN |  |
| Arm 6 | 15mg CBD + 15mg CBN + 5mg CBC |  |

For 5 weeks following study initiation (1 week prior to receipt of product [baseline week], and 4 weeks after receiving product), participants were asked to complete online surveys, sent via email or text. At the end of each week throughout the full study period, participants received a health survey asking them to report their sleep disturbance and sleep quantity using patient reported outcome measures (see below for details). During the baseline week, participants received daily surveys asking them to report their sleep quantity in the previous night. In the subsequent 4 weeks, after receiving their product, participants received a brief survey twice a week asking them to report their product usage and sleep quantity in the previous night. In every study survey following receipt of their product, participants were also prompted to report side effects, and were encouraged to contact the research team directly if at any point they experienced side effects.

The study was registered using ClinicalTrials.gov identifier NCT05552898. Sterling Review Board (IRB) approved the study [Identification number: 9760-EKPauli].

### Randomization

Participants were randomly assigned to one of the six study product arms by a study author (E.K.P.), with an equal chance of being assigned to each group (1:1:1:1:1:1 ratio). Prior to randomization, participants were stratified by their sex at birth (male, female) and baseline self-reported sleep quality in the past week (5 strata, based on response to the following prompt: ‘In the past 7 days, my sleep quality was [ ]’: 1 = ‘Very poor’; 2 = ‘Poor’; 3 = ‘Fair’; 4 = ‘Good’; 5 = ‘Very good’). They were then randomized using a random number generator to one of the study arms using block randomization.

### Outcomes

<u>Patient-Reported Outcomes Measurement Information System (PROMIS™) Sleep Disturbance SF 8A</u> is an 8-item assessment of sleep disturbance during the past 7 days. The items assess characteristics of sleep, frequency of sleep disturbances and sleep quality using a 5-point Likert scale. Cumulative scale scores range from 8 to 40, with higher scores signifying lower sleep quality.^42^

The primary outcomes were the rate of change in the PROMIS Sleep Disturbance 8A scale as well as achieving a minimal clinically important difference (MCID). MCID was defined as a reduction which is greater than or equal to one-half the standard deviation of the baseline score^43^; the MCID standard deviation criterion was calculated by study arm.

<u>Safety</u> The frequency of spontaneously reported side effects and their severity were assessed. Severity was determined based on reported utilization of medical services in response to the side effects according to the following grading schema based on the Common Terminology Criteria for Adverse Events (CTCAE; v5.0 USDHHS): mild: no intervention (medication or medical advice) needed; moderate: a medication was taken due to the side effect; severe: the side effect was significant enough that the participant sought medical care from a healthcare provider, outpatient clinic, or emergency department/room; life threatening: the participant was hospitalized due the side effect.

### Covariates

For precision, we adjusted for baseline demographics, including age, race (Black, multi-racial, some other race, White, prefer not to say), ethnicity (Hispanic, non-Hispanic), sex at birth (male, female) and body mass index (BMI; calculated though self-reported height and weight).

### Power analysis

A power analysis was conducted to ensure sufficient power to detect a significant difference in the change in the PROMIS Sleep Disturbance 8A scale in each study product arm relative to the melatonin isolate arm. A sample size of 164 for each study group would yield 95% power to find a difference in mean change between each study product arm versus the melatonin isolate arm at a two-sided p-value of 0.05 corrected for multiple comparisons (Bonferroni). Recruiting 300 participants per study arm would allow us to maintain adequate sample size under anticipated attrition levels (45%).

### Multiple imputation

The data were imputed 200 times which resulted in 201 versions of the data, including the original, being used for the analyses. A seed was set before conducting the imputations for reproducibility. The chained equations were conducted by study arm and shared the independent variables of sex, age, race, and ethnicity. The imputed variables were BMI and the PROMIS Sleep Disturbance 8a scale. A linear regression model, with the inclusion of height, was used to impute BMI. For the PROMIS Sleep Disturbance 8a scale, a truncated regression with a lower limit of 8 and an upper limit of 40 was used; the lower and upper limits were set based on the minimum and maximum values possible for the scale.

### Statistical analysis

A linear mixed-effects regression model was used to assess the differences in the rate of change in sleep disturbance as measured by the PROMIS Sleep Disturbance 8a scale between each active product arm and CBD isolate. The model was fit using an unstructured covariance matrix with a random-intercept at the individual level and a random-slope at the study week level. The model tested the interaction between product arm and study week, controlling for sex, age, race, ethnicity, and BMI. A logistic regression model was used to assess the differences in the odds of achieving a MCID in their PROMIS Sleep Disturbance 8a score. The logistic regression model tested for differences in odds of MCID between each active product arm and CBD isolate, controlling for sex, age, race, ethnicity, and BMI.

We also conducted three additional (secondary) but *a priori* analyses using linear mixed-effects regression models: (1) an analysis comparing the rate of change in individual PROMIS Sleep Disturbance 8a score items between each active product arm and CBD isolate, (2) an analysis comparing the CBD combination formulations that did *not* contain melatonin to each other, and (3) an analysis comparing formulations containing melatonin to each other. To limit Type 1 error, we used a hierarchical approach in which we first ran an omnibus test to assess overall differences by study arm and, if differences were detected, running pairwise comparisons.

### Software

The Python programming language, version 3.95, and the pandas, version 1.4.3, and numpy, version 1.20.2, were used for data processing. Stata MP (4-core) version 17.0 was used to conduct multiple imputation and the statistical analyses.

## RESULTS

Between March 23 and April 8, 2022, 4,199 individuals were screened for eligibility. Of those screened, 1,793 individuals were eligible, consented to participate and were randomized to a study arm. Between 71% and 74% of participants in each arm completed at least one follow-up survey (see survey completion by study arm in **Appendix Table A**). At the end of the study, 495 participants were excluded from analysis due to completely missing outcome data (classified as ‘no-shows’; i.e., they did not complete *any* surveys asking about their sleep disturbance), leaving 1,298 participants in the final analysis sample (see CONSORT diagram **Figure 1**). For these remaining participants, chained multiple imputation was used to address any missingness. No significant differences in the percentage of ‘no-shows’ were observed between study arms (Pearson χ^2^(5) = 2.082, *p-value=0*.*838*; see **Appendix Table A**).

The mean age of participants at baseline was approximately 46.5 years (standard deviation: 11.5). The majority were female (57%), White (83%) and did not identify as Hispanic (91%). Most participants were either overweight (31%) or obese (43%). About half (50%) had completed at least a bachelors or associates degree and most (63%) were employed. At baseline, approximately 37%, 23%, 32% and 8% of participants had slight, mild, moderate and severe sleep disturbance, respectively, according to their baseline PROMIS Sleep Disturbance 8a score. There were no significant differences in baseline demographic or health characteristics between study arms (see **Table 2**).

**TABLE 2:**
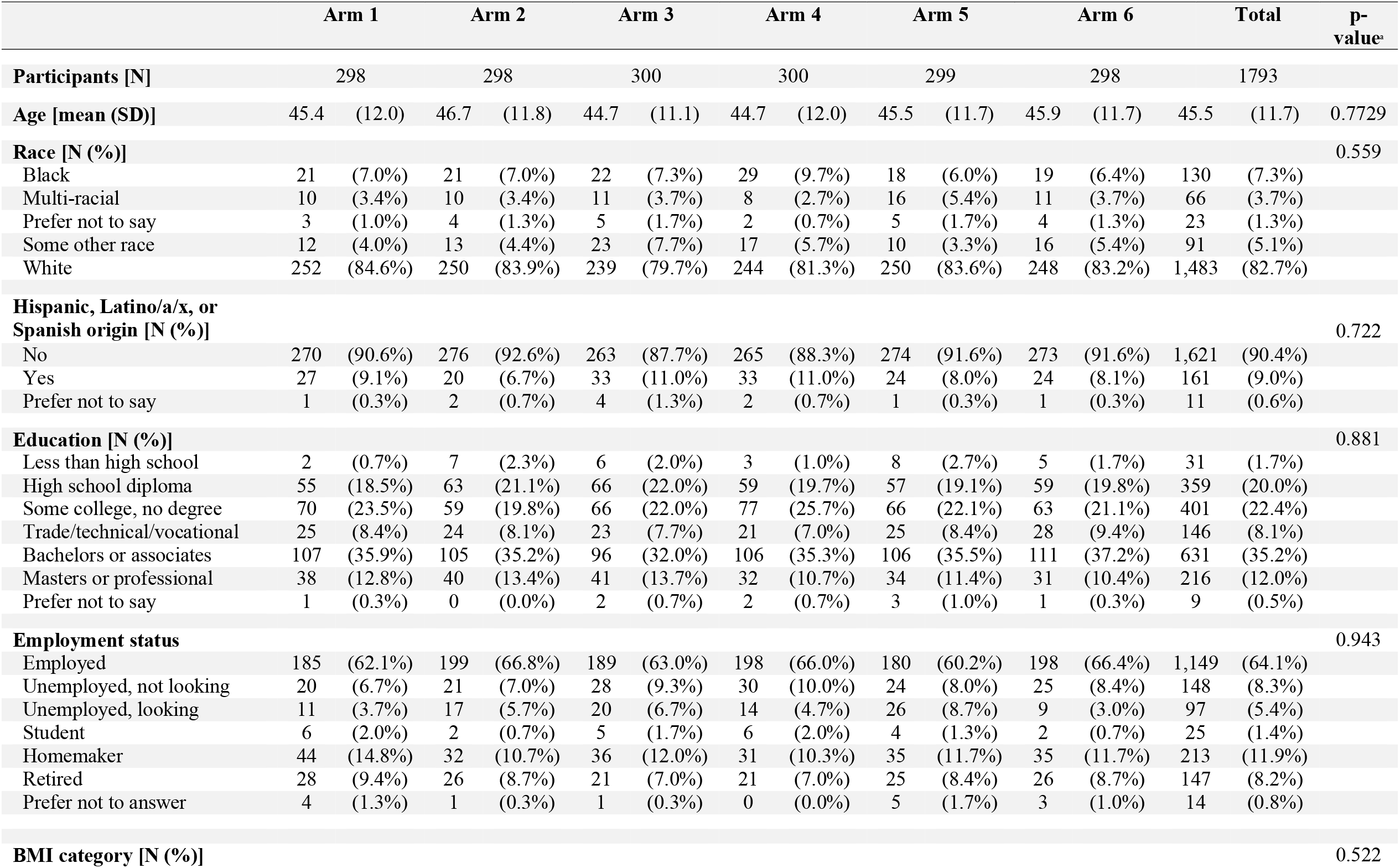
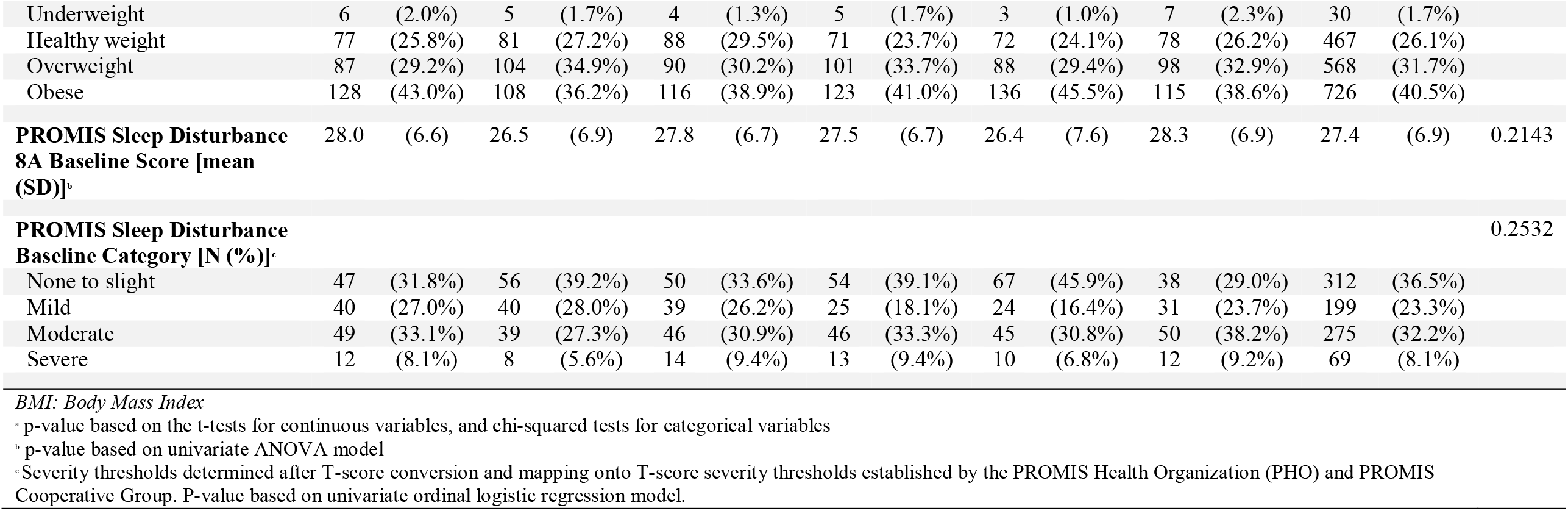
Demographic characteristics of study participants in the Radicle DiscoveryTM Sleep study.

### Product usage

Average daily capsule intake escalated across all study arms, increasing from 1.36 - 1.44 in Week 3 to 1.40 - 1.64 in Week 5, and there were no differences between the study arms (χ2(5)=3.40, *p-value*=0.6386). Throughout the study period, participants in non-melatonin Arms 2, 3, 5 and 6 escalated their average daily capsule intake by 42%, 39%, 45% and 37%, respectively. By contrast, participants assigned to the melatonin-containing Arms 1 (melatonin combination) and 4 (melatonin isolate) escalated their average daily capsule intake by 32% and 33%, respectively.

### Change in PROMIS Sleep Disturbance 8a score

Within the study timeframe, all study arms experienced a significant reduction in the PROMIS Sleep Disturbance 8a score from baseline (within-group *p* < 0.0001; see **Table 4**). Overall, there were significant differences in the rate of change between study arms (*F*[5,9414.2] = 2.29, *p* = 0.0433).

Compared to Arm 3 (15 mg CBD isolate), Arm 1 (melatonin combination) exhibited greater reductions in sleep disturbance score, though these effects were only moderately significant (*Coef*. = -0.4, 95% CI [-0.8, 0.0], *p* = 0.059). No other differences were observed between any other active arm and Arm 3 (see **Table 3**).

**TABLE 3:** Mean difference (MD) in PROMIS Sleep Disturbance 8a scores from baseline to study conclusion, based on adjusted linear mixed effects regression model.

| Study arm#Week<sup>b</sup> | MD (SE) | Statistics | 95% CI |
| --- | --- | --- | --- |
| Arm 1 | -0.4 (0.2) | $t = -1.89, p < 0.059$ | (-0.8, 0) |
| Arm 2 | 0.1 (0.2) | $t = 0.62, p < 0.538$ | (-0.3, 0.5) |
| Arm 4 | -0.3 (0.2) | $t = -1.54, p < 0.123$ | (-0.7, 0.1) |
| Arm 5 | 0.1 (0.2) | $t = 0.64, p < 0.525$ | (-0.3, 0.5) |
| Arm 6 | -0.1 (0.2) | $t = -0.27, p < 0.786$ | (-0.5, 0.4) |

*SE: standard error of the estimate; CI: Confidence interval of the estimate*
Multiple imputed linear mixed-effects regression model with random intercept at the individual level and random slope at week level; $F(20, 102788.8) = 46.56, p\text{-value} < 0.0001$
<sup>a</sup> Adjusting for sex, age, race, ethnicity and body mass index
<sup>b</sup> Joint-test of main effects significant; $F(5, 9414.2) = 2.92, p\text{-value} = 0.0433$

**TABLE 4:**
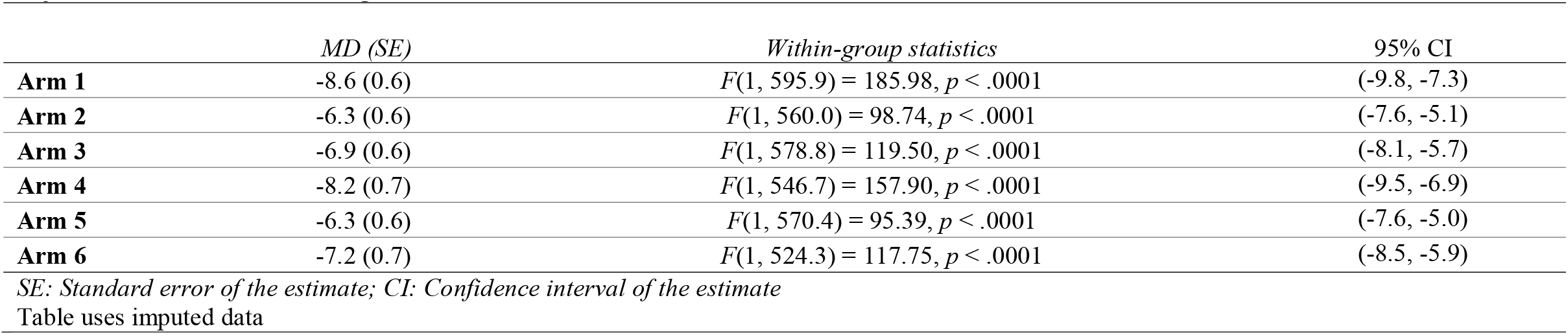
Within-group mean difference (MD) in PROMIS Sleep Disturbance 8a scores from baseline to study conclusion, based on adjusted linear mixed effects regression model.

|  | MD (SE) | Within-group statistics | 95% CI |
| --- | --- | --- | --- |
| Arm 1 | -8.6 (0.6) | $F(1, 595.9) = 185.98, p < .0001$ | (-9.8, -7.3) |
| Arm 2 | -6.3 (0.6) | $F(1, 560.0) = 98.74, p < .0001$ | (-7.6, -5.1) |
| Arm 3 | -6.9 (0.6) | $F(1, 578.8) = 119.50, p < .0001$ | (-8.1, -5.7) |
| Arm 4 | -8.2 (0.7) | $F(1, 546.7) = 157.90, p < .0001$ | (-9.5, -6.9) |
| Arm 5 | -6.3 (0.6) | $F(1, 570.4) = 95.39, p < .0001$ | (-7.6, -5.0) |
| Arm 6 | -7.2 (0.7) | $F(1, 524.3) = 117.75, p < .0001$ | (-8.5, -5.9) |

*SE: Standard error of the estimate; CI: Confidence interval of the estimate*
Table uses imputed data

When assessing rate of change in score by each PROMIS Sleep Disturbance 8a scale question separately, we observed that those in Arm 1 exhibited greater reductions in sleep disturbance relative to Arm 3 across the following items: ‘My sleep was refreshing’ (*Coef*.*=* -0.1, 95% CI [-0.1, 0.0], *p* = 0.049), ‘I had a problem with my sleep’ (*Coef*.*=* -0.1, 95% CI [-0.2, 0.0], *p* = 0.005), and ‘My sleep was restless’ (*Coef*.*=* -0.1, 95% CI [-0.1, 0.0], *p* = 0.019). No other differences were observed between any other active arm and Arm 3 for these items. No differences were observed between any active arm and Arm 3 for the items ‘My sleep quality was .’, ‘I had difficulty falling asleep’, ‘I tried hard to get to sleep’, ‘I was worried about not being able to fall asleep’, and ‘I was satisfied with my sleep’ (see **Table 5**).

**Table 5:**
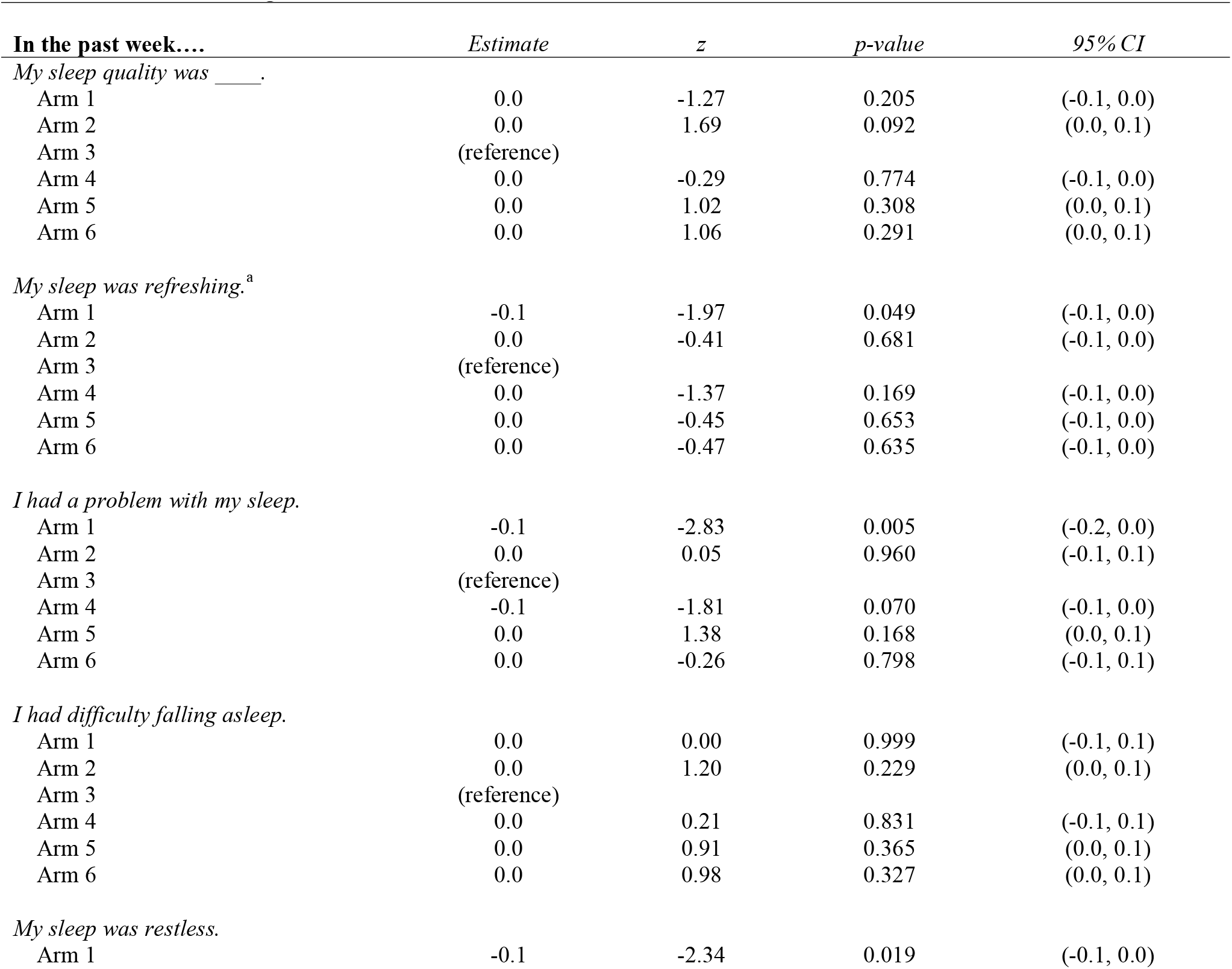
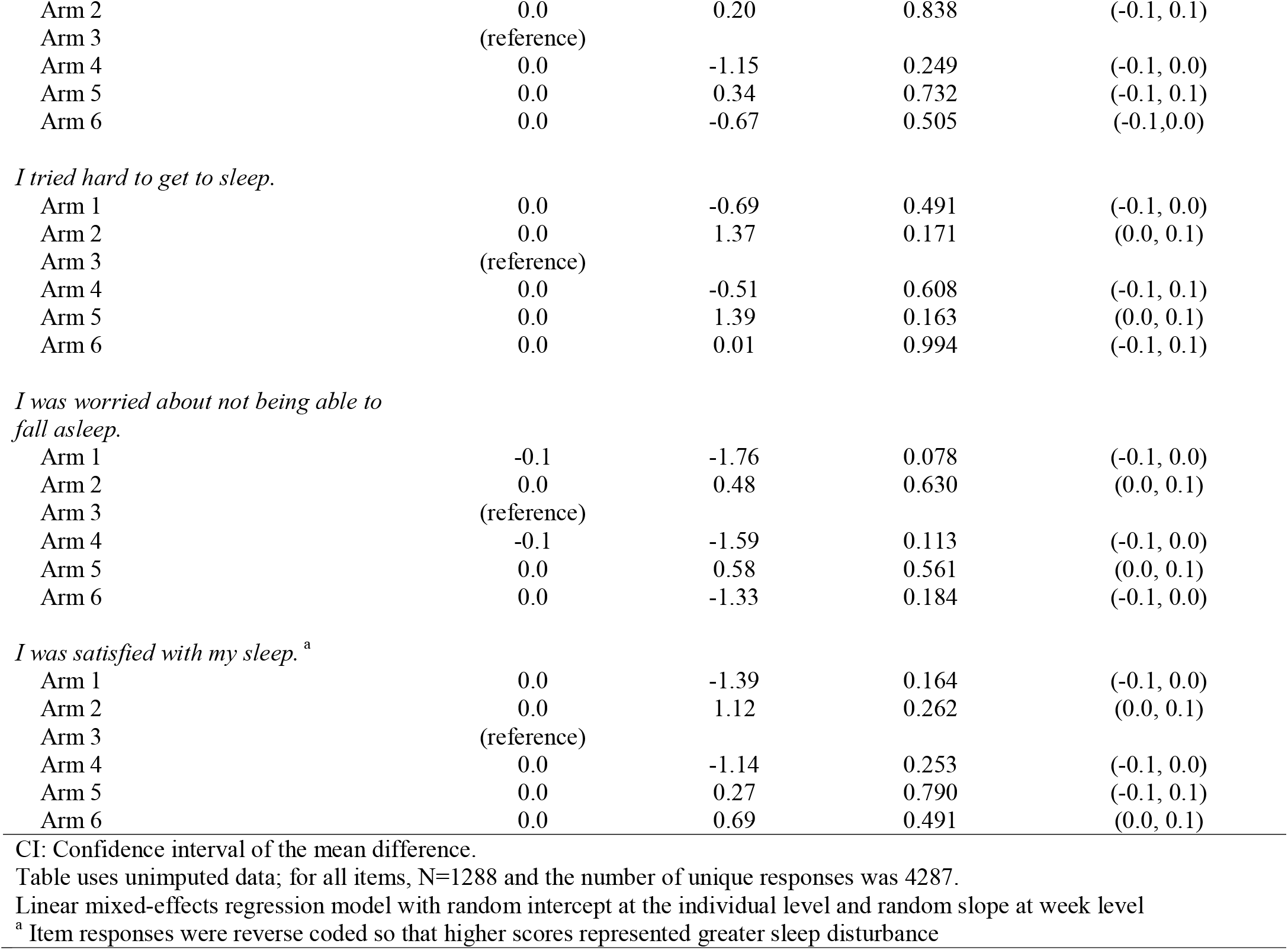
Mean difference (MD) in PROMIS Sleep Disturbance SF 8a item score from baseline to study conclusion, based on linear mixed effects regression model.

| **In the past week....** | *Estimate* | *z* | *p-value* | *95% CI* |
| --- | --- | --- | --- | --- |
| *My sleep quality was ____.* |  |  |  |  |
| Arm 1 | 0.0 | -1.27 | 0.205 | (-0.1, 0.0) |
| Arm 2 | 0.0 | 1.69 | 0.092 | (0.0, 0.1) |
| Arm 3 | (reference) |  |  |  |
| Arm 4 | 0.0 | -0.29 | 0.774 | (-0.1, 0.0) |
| Arm 5 | 0.0 | 1.02 | 0.308 | (0.0, 0.1) |
| Arm 6 | 0.0 | 1.06 | 0.291 | (0.0, 0.1) |
| *My sleep was refreshing.<sup>a</sup>* |  |  |  |  |
| Arm 1 | -0.1 | -1.97 | 0.049 | (-0.1, 0.0) |
| Arm 2 | 0.0 | -0.41 | 0.681 | (-0.1, 0.0) |
| Arm 3 | (reference) |  |  |  |
| Arm 4 | 0.0 | -1.37 | 0.169 | (-0.1, 0.0) |
| Arm 5 | 0.0 | -0.45 | 0.653 | (-0.1, 0.0) |
| Arm 6 | 0.0 | -0.47 | 0.635 | (-0.1, 0.0) |
| *I had a problem with my sleep.* |  |  |  |  |
| Arm 1 | -0.1 | -2.83 | 0.005 | (-0.2, 0.0) |
| Arm 2 | 0.0 | 0.05 | 0.960 | (-0.1, 0.1) |
| Arm 3 | (reference) |  |  |  |
| Arm 4 | -0.1 | -1.81 | 0.070 | (-0.1, 0.0) |
| Arm 5 | 0.0 | 1.38 | 0.168 | (0.0, 0.1) |
| Arm 6 | 0.0 | -0.26 | 0.798 | (-0.1, 0.1) |
| *I had difficulty falling asleep.* |  |  |  |  |
| Arm 1 | 0.0 | 0.00 | 0.999 | (-0.1, 0.1) |
| Arm 2 | 0.0 | 1.20 | 0.229 | (0.0, 0.1) |
| Arm 3 | (reference) |  |  |  |
| Arm 4 | 0.0 | 0.21 | 0.831 | (-0.1, 0.1) |
| Arm 5 | 0.0 | 0.91 | 0.365 | (0.0, 0.1) |
| Arm 6 | 0.0 | 0.98 | 0.327 | (0.0, 0.1) |
| *My sleep was restless.* |  |  |  |  |
| Arm 1 | -0.1 | -2.34 | 0.019 | (-0.1, 0.0) |
| Arm 2 | 0.0 | 0.20 | 0.838 | (-0.1, 0.1) |
| Arm 3 | (reference) |  |  |  |
| Arm 4 | 0.0 | -1.15 | 0.249 | (-0.1, 0.0) |
| Arm 5 | 0.0 | 0.34 | 0.732 | (-0.1, 0.1) |
| Arm 6 | 0.0 | -0.67 | 0.505 | (-0.1,0.0) |
| *I tried hard to get to sleep.* |  |  |  |  |
| Arm 1 | 0.0 | -0.69 | 0.491 | (-0.1, 0.0) |
| Arm 2 | 0.0 | 1.37 | 0.171 | (0.0, 0.1) |
| Arm 3 | (reference) |  |  |  |
| Arm 4 | 0.0 | -0.51 | 0.608 | (-0.1, 0.1) |
| Arm 5 | 0.0 | 1.39 | 0.163 | (0.0, 0.1) |
| Arm 6 | 0.0 | 0.01 | 0.994 | (-0.1, 0.1) |
| *I was worried about not being able to fall asleep.* |  |  |  |  |
| Arm 1 | -0.1 | -1.76 | 0.078 | (-0.1, 0.0) |
| Arm 2 | 0.0 | 0.48 | 0.630 | (0.0, 0.1) |
| Arm 3 | (reference) |  |  |  |
| Arm 4 | -0.1 | -1.59 | 0.113 | (-0.1, 0.0) |
| Arm 5 | 0.0 | 0.58 | 0.561 | (0.0, 0.1) |
| Arm 6 | 0.0 | -1.33 | 0.184 | (-0.1, 0.0) |
| *I was satisfied with my sleep.*<sup>a</sup> |  |  |  |  |
| Arm 1 | 0.0 | -1.39 | 0.164 | (-0.1, 0.0) |
| Arm 2 | 0.0 | 1.12 | 0.262 | (0.0, 0.1) |
| Arm 3 | (reference) |  |  |  |
| Arm 4 | 0.0 | -1.14 | 0.253 | (-0.1, 0.0) |
| Arm 5 | 0.0 | 0.27 | 0.790 | (-0.1, 0.1) |
| Arm 6 | 0.0 | 0.69 | 0.491 | (0.0, 0.1) |

CI: Confidence interval of the mean difference.
Table uses unimputed data; for all items, N=1288 and the number of unique responses was 4287.
Linear mixed-effects regression model with random intercept at the individual level and random slope at week level
<sup>a</sup> Item responses were reverse coded so that higher scores represented greater sleep disturbance

We also found no overall differences in the rate of change of overall sleep disturbance score between any combination CBD formulation containing no melatonin (Arms 2, Arm 5 and Arm 6; F[3,5585.6] = 0.40, *p* = 0.7504). Further, we observed no differences in the rate of change of overall sleep disturbance score between the formulations containing melatonin (Arms 1 vs. 4; *Coef*. = -0.07, 95% CI [-0.48, 0.34], *p* = 0.734).

### Achievement of MCID

Across all study arms, 67% of individuals experienced a MCID in their sleep disturbance score. Of the arms which did not contain melatonin, 56.4% of Arm 2, 68.4% of Arm 3, 60.0% of Arm 5, and 72.1% of Arm 6 experienced a MCID. Of the arms containing melatonin, 71.0% of those assigned to Arm 4 (melatonin only) experienced a MCID compared to 75.3% of those in Arm 1 (melatonin combination). Overall, there was a significant difference between arms in the odds of experiencing a MCID (F[5, 9414.2]=2.29, p-value=0.0433). However, no significant differences in the odds of achieving MCID were observed between Arm 3 and any other active study arm (see **Table 6)**.

**TABLE 6:** Adjusted^a^ odds ratio (aOR) of achieving a minimal clinically important difference in PROMIS Sleep Disturbance 8a scores.

|  |  | *aOR* | *95% CI* |
| --- | --- | --- | --- |
| **Study arm<sup>b</sup>** |  |  |  |
|  | **Arm 1** | 1.4 | (0.8, 2.4) |
|  | **Arm 2** | 0.7 | (0.4, 1.2) |
|  | **Arm 3** | (reference) |  |
|  | **Arm 4** | 1.3 | (0.8, 2.2) |
|  | **Arm 5** | 0.8 | (0.5, 1.4) |
|  | **Arm 6** | 1.1 | (0.7, 1.8) |

*CI: Confidence interval of the estimate*
Multiple imputed logistic regression; F(14, 22175.9)=1.02, p-value=0.4272
<sup>a</sup> Adjusted for sex, age, race, ethnicity and body mass index
<sup>b</sup> Joint-test of main effects significant; F(5, 9414.2)=2.29, p-value=0.0433

In secondary analyses, we found no overall differences in odds of experiencing a MCID between any of the formulations containing no melatonin (Arms 2, Arm 5 and Arm 6; F [3,4281.5] = 1.14; *p* = 0.3307). We also observed no differences between the formulations containing melatonin (Arms 1 and 4; *Coef*. = 0.06, 95% CI [-0.49, 0.60], *p* = 0.837).

### Safety

Approximately 11.9% (n=199) of participants reported experiencing at least one side effect during the study. About 85% of side effects reported during study follow-up were classified as mild, and 14% were classified as moderate (1% failed to provide information regarding side effect severity). No reported side effects were classified as severe or life-threatening. The most common reported side effects included fatigue/grogginess (2.7%; n=49), insomnia/sleep disturbance (1.4%; n=26), headache (1.1%; n=19), upset stomach (1.0%; n=18), dry mouth (0.8%; n=14), nausea/vomiting (0.8%; n=14), restless feeling (0.8%; n=14), vivid dreams or nightmares (0.8%; n=14). The percentage of participants experiencing at least one side effect was similar across study arms (ranging from 9.6% to 13.7%; see **Appendix Table B**).

## DISCUSSION

In this randomized, double-blinded controlled trial to evaluate the effects of different orally ingested cannabinoid and melatonin formulations on sleep disturbance, we observed that a 15mg of CBD reduced self-reported sleep disturbance over the course of 4 weeks. The addition of minor cannabinoids (15mg CBN, alone or in combination with 5 mg CBC) did not impact the therapeutic effects of 15 mg CBD. Moreover, we found no evidence of a difference in effect on overall sleep disturbance score between 15mg CBD isolate and formulations containing 5 mg melatonin, alone or in combination with 15mg CBD and 15mg CBN. However, when examining changes in each PROMIS Sleep Disturbance 8a scale question score separately, we observed that those taking the 5mg melatonin in combination with 15mg CBD and 15mg CBN reported greater improvements in the restless and refreshing aspects of their sleep relative to those taking CBD isolate, though changes in self-reported sleep quality, sleep satisfaction, and in difficulties and worries over falling asleep did not vary between any formulation relative to CBD isolate. Secondary analyses also revealed that the addition of 15mg CBD and 15mg CBN did not significantly impact the therapeutic effects of a formulation containing 5mg melatonin on overall sleep disturbance score. Notably, all study arms led to significant improvements in sleep disturbance and exhibited favorable safety profiles.

Few clinical studies have examined the effect of CBD, with or without the addition of minor cannabinoids, for the improvement of sleep. This study is among the first to evaluate the safety and effectiveness of CBD dose ranges and formulations commonly found within commercially available CBD products. Our results demonstrate that the relatively lower doses of CBD found within these orally ingested products may be safe for chronic use and sufficient to produce significant improvements in symptoms of sleep disturbance. These effects, however, do not exceed that of 5 mg melatonin. Moreover, we found no evidence that the low doses of CBN and CBC can improve the effect of formulations containing low doses of CBD or melatonin isolate.

Our findings regarding the lack of additional therapeutic effect from the addition of CBN are noteworthy as cannabis product manufacturers have recently begun to tout the sleep-inducing effects of CBN,^44^ though preclinical and clinical research in support of these claims is scarce and outdated.^36^ Indeed, our study represents the first clinical trial to evaluate the use of CBN for sleep using validated sleep measures. Our findings suggest that 15mg of CBN may confer little added benefit to a sleep aid product. We note, however, that our findings reflect a relatively lower dosage of orally ingested CBN and may not be generalizable to higher dosages or other modes of administration of the cannabinoid.

In our secondary analyses, we did not find evidence that the addition of CBC to formulations containing CBD and CBN will impact their therapeutic effect on sleep quality. We did, however, observe substantially higher likelihood of experiencing a MCID among those in the CBD combination arm containing CBC (72%) compared to the arms containing just CBD and CBN (56% and 60% in the Full and Isolate spectrum CBD combination arms, respectively), suggesting CBC might impart some effect on sleep improvement. However, as this was not a planned comparison for the current study, and as the omnibus test between the CBD combination arms did not reach statistical significance, we are unable to fully interpret the meaning of this trend. Thus, further studies are needed to thoroughly characterize the impacts of CBC on sleep.

Preclinical research suggests that certain cannabinoids and other components of the *Cannabis* plant could work synergistically, stimulating a greater effect than that if CBD or Δ9-THC were examined in isolation – a phenomenon known as the ‘entourage effect’.^45^ As we observed no difference in effect between the CBD isolate and cannabinoid combination formulations, we found no evidence of an entourage effect with CBD, CBC, and CBN in these trial products. However, these findings may only be generalized to the specific dosages of these cannabinoids in this sample. We cannot exclude, from the present evidence, the possibility that higher concentrations of these minor cannabinoids could modulate the effects of CBD.

We observed that participants assigned to arms without melatonin averaged a higher daily capsule intake compared to participants taking melatonin-containing capsules. Importantly, we did not detect an increase in safety concerns in arms with a higher average daily intake. Although not statistically significant, this trend may suggest that without melatonin, higher doses of cannabinoids may be necessary to induce the desired effects on sleep quality. Moreover, it is possible that melatonin capsule dose was more optimally calibrated relative to the doses of cannabinoids.

Previous clinical research on melatonin for insomnia and other common sleep disorders suggests that its effects are modest and inferior to prescription medications.^46^ As we did not find any significant differences between CBD isolate and the melatonin or CBD combination formulations, such conclusions regarding modest relative effects could be extended to the formulations in this sample. Nonetheless, as melatonin and CBD both demonstrate a highly favorable safety and tolerability profile, these alternative therapies could still play a role in the treatment of common sleep disorders, especially given the harmful side effects of common pharmacological treatments for these disorders.^8^

This study has several limitations. First, about 28% of participants did not complete any follow-up surveys and were therefore excluded from the study. While our overall attrition levels still fell below anticipated attrition levels of 45% and the study was adequately powered to detect significant sleep changes, differential loss to follow-up could induce post-randomization confounding.^47^ As those who were excluded did not fill out any surveys beyond the screener, we were limited in our ability to evaluate characteristics of those excluded and assess potential imbalances across study arms. We were also unable to run a sensitivity analysis including the excluded individuals as they did not provide any PROMIS Sleep Disturbance 8a scores and imputing their data would not be appropriate as their observations were determined to be missing not at random (MNAR; as no study period surveys were completed by these individuals, the missingness is thus dependent on unobserved data). Nonetheless, we did not find any significant differences in the percentage of excluded individuals between study arms, and there were no significant differences in baseline demographic or health characteristics between study arms in the final sample, indicating that balance was maintained across study arms despite changes to the study sample post-randomization.

Additionally, as there was no placebo control within this study, we cannot determine if and how much the observed effects may be due to participant expectations/placebo. In previous clinical research, melatonin has been shown to have modest effects on sleep relative to placebo,^48^ though clinical evidence of CBD’s effect relative to placebo remains limited, albeit promising.^23–25^ Notably, previous clinical research suggests that placebo response could play a major role in the effect of CBD on stress and anxiety,^49^ though the impact of this response has yet to be explored for CBD and sleep. Further placebo-controlled studies are needed to determine the therapeutic effects of CBD for sleep.

We see this data as “real world” data, as it was collected from a population that was using the products in a manner and setting like that of real consumers of these products. Without exhaustive eligibility criteria and intensive monitoring, the missingness and heterogeneity of the data may be greater than that of traditional clinical trials. However, many traditional clinical trials have limited external validity because the characteristics and behaviors of participants may not reflect those of real-world users. As such, real world studies have unique value in their ability to provide complementary evidence to support clinical trial designs and help guide regulatory and clinical decisions.^50,51^

Our study is the first randomized, blinded, controlled trial which compares the effects of CBD and melatonin on sleep, and further investigates therapeutic benefits of combining CBD isolate with melatonin or minor cannabinoids. Our findings represent an essential scientific advancement towards thoroughly characterizing and contrasting the effects of commonly used non-prescription sleep disorder treatments.

## Data Availability

All data produced in the present study are available upon reasonable request to the authors

---

**Appendix Table A:**
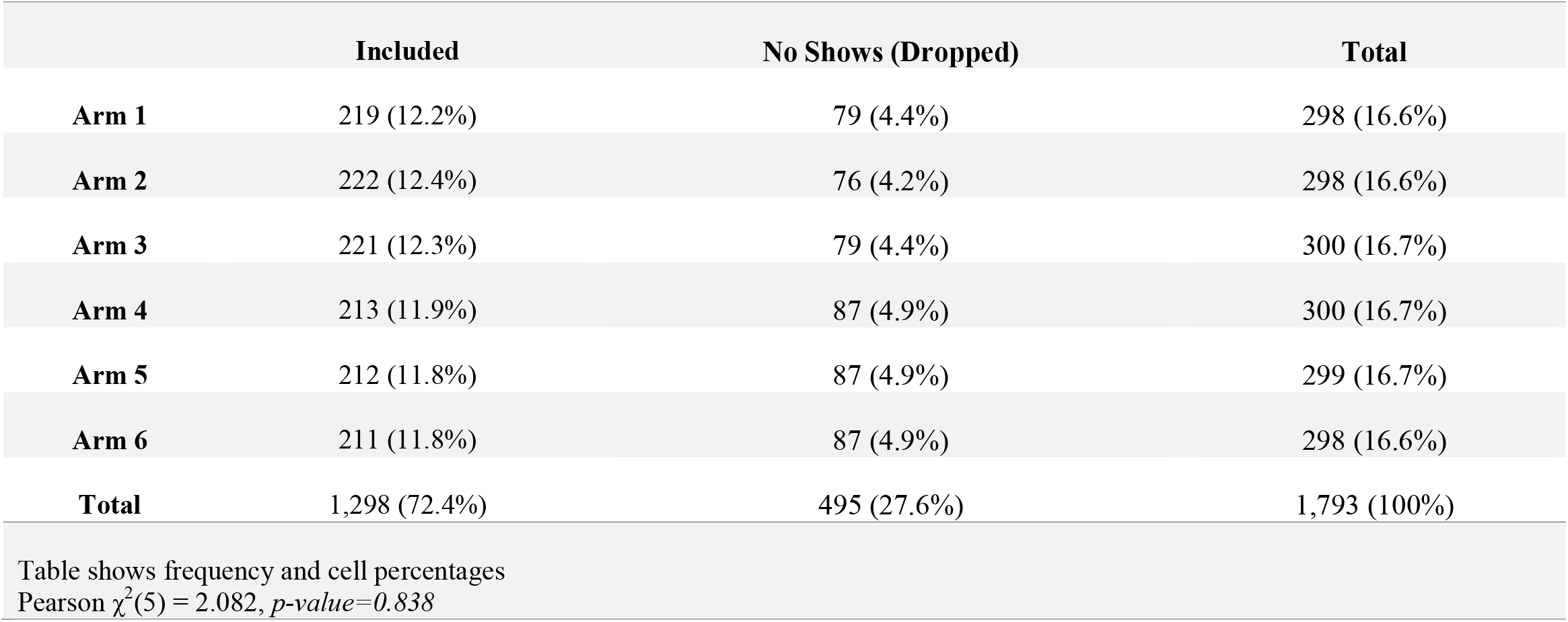
Survey completion by study arm.

**Appendix Table B:** Side effects reported by study arm.

|  | Arm 1 | Arm 2 | Arm 3 | Arm 4 | Arm 5 | Arm 6 | Total |
| --- | --- | --- | --- | --- | --- | --- | --- |
| Fatigue/grogginess [N] | 9 | 5 | 8 | 12 | 8 | 7 | 49 |
| Insomnia/sleep disturbance [N] | 2 | 7 | 5 | 4 | 4 | 4 | 26 |
| Headaches [N] | 3 | 4 | 5 | 3 | 2 | 2 | 19 |
| Upset stomach [N] | 3 | 0 | 6 | 5 | 1 | 3 | 18 |
| Dry mouth [N] | 1 | 3 | 2 | 2 | 5 | 1 | 14 |
| Nausea / vomiting [N] | 3 | 2 | 1 | 4 | 2 | 2 | 14 |
| Restless feeling [N] | 1 | 1 | 5 | 1 | 4 | 2 | 14 |
| Vivid dreams / nightmares [N] | 3 | 1 | 3 | 3 | 2 | 2 | 14 |
| Heartburn [N] | 1 | 0 | 2 | 5 | 1 | 1 | 10 |
| Diarrhea [N] | 1 | 1 | 1 | 1 | 1 | 0 | 5 |
| Constipation [N] | 2 | 0 | 1 | 1 | 0 | 1 | 5 |
| Dizziness [N] | 0 | 0 | 1 | 1 | 1 | 1 | 4 |
| Food cravings [N] | 0 | 1 | 0 | 1 | 2 | 0 | 4 |
| Itching [N] | 0 | 0 | 1 | 0 | 2 | 0 | 3 |
| Anxiety [N] | 0 | 2 | 1 | 0 | 0 | 0 | 3 |
| Irritability [N] | 0 | 0 | 1 | 0 | 0 | 1 | 2 |
| Mental fog [N] | 0 | 1 | 0 | 0 | 0 | 1 | 2 |
| Gas [N] | 0 | 0 | 1 | 0 | 1 | 0 | 2 |
| Weight gain [N] | 1 | 0 | 0 | 0 | 0 | 1 | 2 |
| Stomach cramps [N] | 1 | 0 | 0 | 1 | 0 | 0 | 2 |
| Sweating [N] | 0 | 0 | 2 | 0 | 0 | 0 | 2 |
| Back pain [N] | 0 | 0 | 0 | 0 | 1 | 0 | 1 |
| Leg cramps [N] | 0 | 0 | 1 | 0 | 0 | 0 | 1 |
| Shoulder pain [N] | 0 | 0 | 0 | 0 | 0 | 1 | 1 |
| Swollen feet [N] | 0 | 0 | 0 | 1 | 0 | 0 | 1 |
| Body aches [N] | 0 | 0 | 1 | 0 | 0 | 0 | 1 |
| Sore throat [N] | 0 | 0 | 1 | 0 | 0 | 0 | 1 |
| Rash [N] | 0 | 0 | 0 | 0 | 1 | 0 | 1 |
| Hot/cold flashes [N] | 0 | 0 | 0 | 0 | 1 | 0 | 1 |
| **Total reporting any side effects [N(%)]** | 30 (10.1%) | 31 (10.4%) | 41 (13.7%) | 36 (12.0%) | 32 (10.7%) | 29 (9.7%) | 199 (11.9%) |

